# Embedded point of care stratified block randomization: demonstration of the point of care randomization (POCR) engine with an electronic health record pragmatic clinical trial

**DOI:** 10.64898/2026.01.26.26344847

**Authors:** Catherine Sarkisian, Khalda Ibrahim, Sitaram Vangala, Chad Wes Villaflores, Eric M. Cheng, William Turner, Richard K. Leuchter, Amy Machado, Julia Tabar, Jade A. Verdeflor, Jory Purvis, Alina Goncharova, Mark J. Pletcher

## Abstract

We describe a new custom feature within our Epic Systems electronic health record (EHR) that automates stratified randomization at the point-of-care or order. As a demonstration use-case, we conducted a randomized trial of a provider-facing alert for short-interval HbA1c orders. Over 3 months the alert dramatically reduced repeat orders. This transportable clinical informatics application transforms health systems’ ability to conduct pragmatic clinical trials and deliver clinical care within the EHR.

## MAIN TEXT

The widespread adoption of electronic health records (EHR) offers tremendous opportunity to improve healthcare delivery and patient outcomes. Accordingly, measuring the impact of EHR interventions is a timely and active area of translational informatics research. Typically, health systems and their clinical informatics and quality improvement teams implement EHR interventions that “make sense” purely on the grounds that the mechanisms by which they could improve outcomes are bioplausible. But these interventions are rarely tested in randomized controlled trials (RCTs) to prove that they are effective and do not cause unintended harms.^1^ Common examples include provider-facing point-of-care decision support alerts and patient-facing health maintenance reminders. While the biomedical literature is full of pre-post analyses of these types of EHR interventions, it is difficult to confirm whether these projects are truly successful or whether pre-post differences represent secular trends and/or regression to the mean.^2^

Until recently, it has not been possible to randomize interventions by patient, provider, or encounter in real time, stratified, and entirely within routine, EHR-based clinical workflows. Some research teams utilize “pseudorandomization” using surrogate methods such as day of week or odd/even medical record number.^3^ Pseudorandomization does not allow for techniques—such as block or stratified randomization—that facilitate balanced assignment within treatment groups; it makes the randomization assignment for any given unit predictable and unblinded; and it may build in structural group differences (e.g., patients who present on Sunday may be different than patients presenting on Monday). True randomization with stratification and blocking currently requires cumbersome processes—for example, randomizing potentially eligible patients or providers outside the EHR with a random number generator and then assigning their study arm prior to an upcoming encounter in which the EHR intervention would be triggered. Workaround processes like these fix arm assignments at the beginning of the intervention (such as when an alert “goes live”), even though enrollment may unfold over months. Someone randomized *ex ante* may never actually trigger the action of interest (e.g., by ordering a certain medication or lab test), and individuals who were not already randomized *ex ante* (e.g., new hires or newly empaneled patients) are absolutely excluded from the study. Current EHRs cannot determine eligibility and randomize assignment *ad hoc* at the point-of-care or point-of-order. True, real-time EHR-based randomization within existing clinical workflows would overcome these barriers, transforming the workflow of pragmatic clinical trials and the delivery of clinical care, marking a critical advancement in the ability to generate high-quality, causal evidence within healthcare systems.

In 2020, the Clinical and Translational Sciences Institute (CTSI) at the University of California, San Francisco (UCSF) developed a new feature to automate randomization within the EHR. The feature, programmed in Caché, uses Epic rule infrastructure to implement inclusion criteria, and generate and use blocks of randomization assignments. Configuration details, randomization assignments, and context information (including stratification variables) are stored in custom master files in Chronicles and exported to Clarity (Epic’s reporting database). The system supports randomization of patients, encounters, clinicians, clinical units, or any other type of “object” in Epic, with programmable randomization ratios, strata, and randomly varying block sizes to ensure balance and unpredictability. UCSF uses this Point-of-Care Randomization (POCR) engine for randomized trials of interventions in their EHR. ^4 5 6^

UCLA, though also part of the larger University of California, operates as an independent health system supported by its own instance of Epic that is separate from UCSF’s instance. UCLA Health has a long-standing commitment to being a learning health system.^7^ As part of this mission, the health system supports UCLA Healthcare Value Analytics and Solutions (UVAS)—a research team based in the School of Medicine that works closely with Chief Medical Information Officer Dr. Eric Cheng and UCLA Informatics Systems and Solutions (ISS) to provide timely analyses to inform health system operations and improve healthcare delivery. In 2023 UVAS, ISS, and the UCLA Office of Health Informatics and Analytics (OHIA) partnered with UCSF to deploy the randomization engine at UCLA Health.

As a demonstration use-case for the randomization engine at UCLA, UVAS collaborated with the Laboratory Stewardship Task Force on an initiative to reduce testing that is unlikely to improve care. We conducted a pragmatic quality improvement RCT of provider-facing clinical decision support (CDS) designed to discourage inpatient orders for hemoglobin A1c (HbA1c) within 30 days of prior HbA1c testing (which we term “short-interval HbA1c orders”); repeating HbA1c testing within 30 days is generally not indicated.^8^ This alert criteria included recently collected specimens with pending results. Through an interdisciplinary iterative design process we developed our intervention, a provider-facing interruptive alert to display the date and results of HbA1c testing performed in the previous 30 days. (See Figure 1). We hypothesized that if busy providers taking care of adult inpatients: 1) were notified in real time that their patient had had HbA1c testing performed recently; 2) were automatically provided with the date and results of that testing; and 3) were advised that repeat testing within 30 days is “generally not recommended,” they would not finalize their initiated HbA1c orders.

**Figure 1.**
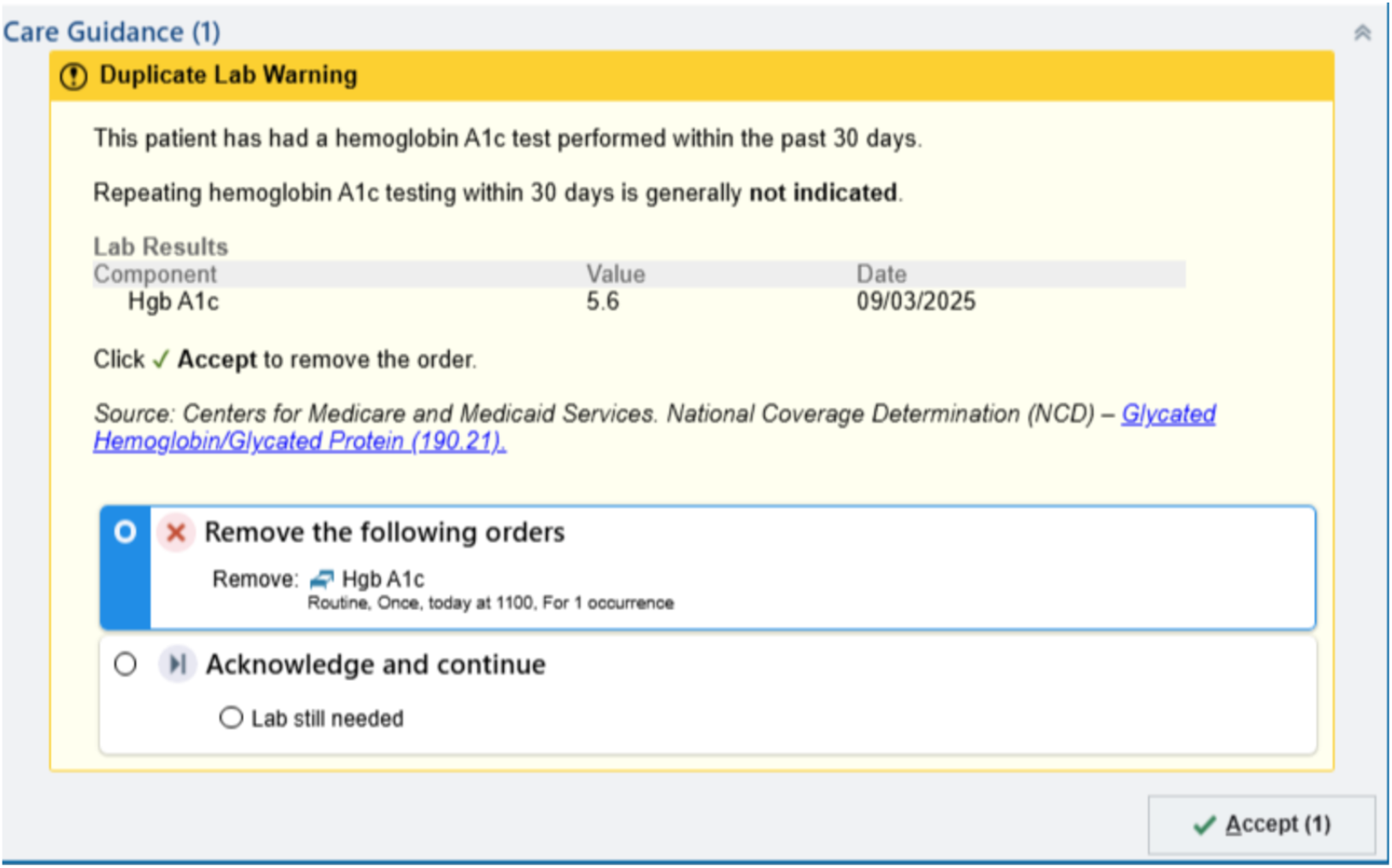
Provider-facing interruptive alert.

The study took place over 3 months (May 12-August 12, 2025). Unlike our usual practice when launching a new informatics alert, we did not deliver system-wide electronic mail education to all providers. Patients were the unit of randomization: eligible patients included all hospitalized patients (inpatient or observation status) over age 18 years for whom a provider was initiating an order for a HbA1c test and whose chart included either HbA1c results from the previous 30 days or a pending HbA1c result. Whenever providers initiated an order for repeat HbA1c testing, eligible patients were silently enrolled and randomized solely within the EHR using the new POCR Engine. A random half of patients’ providers were immediately shown the interruptive alert (intervention arm); the other half were silently enrolled with no alert fired (control arm—care as usual). The randomization within each arm was stratified based on patient age (18-55, 55+) and most recent HbA1c value (<6.5%, 6.5-8%, >8%). We randomly generated blocks of 4, 6 or 8 patients within each strata.

During the study period, 333 patients were enrolled and randomized, with 172 in the intervention arm and 161 in the control arm. (See Figure 2). Patient characteristics are shown in Table 1. Mean age was 62.2 years, with 36.0% female. Self-identified race/ethnicity was: 10.8% Asian, 13.2% Black or African American, 27.0% Latino/Hispanic, 35.7% White.

**Table 1.**
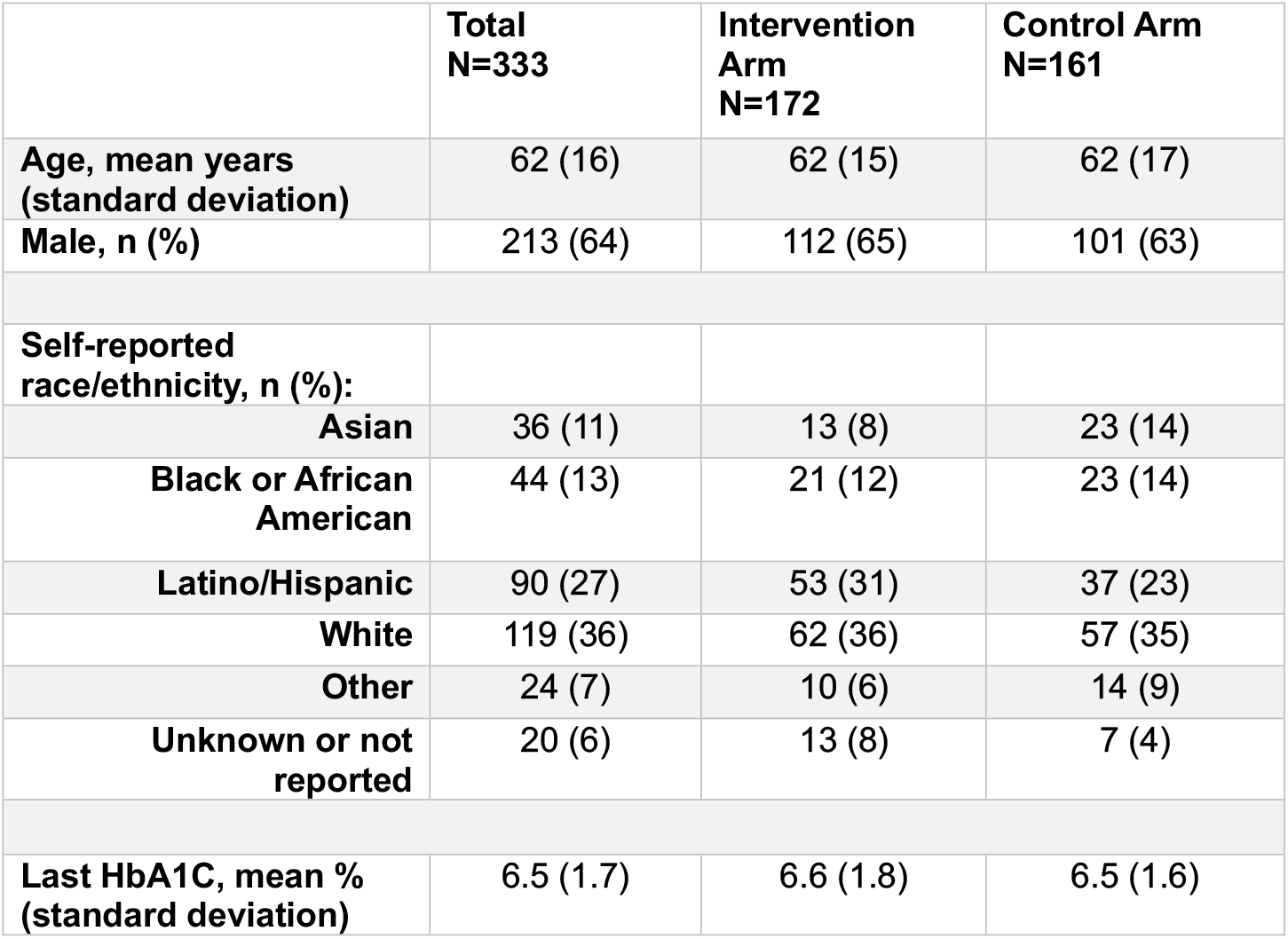
Patient Characteristics.

**Fig. 2.**
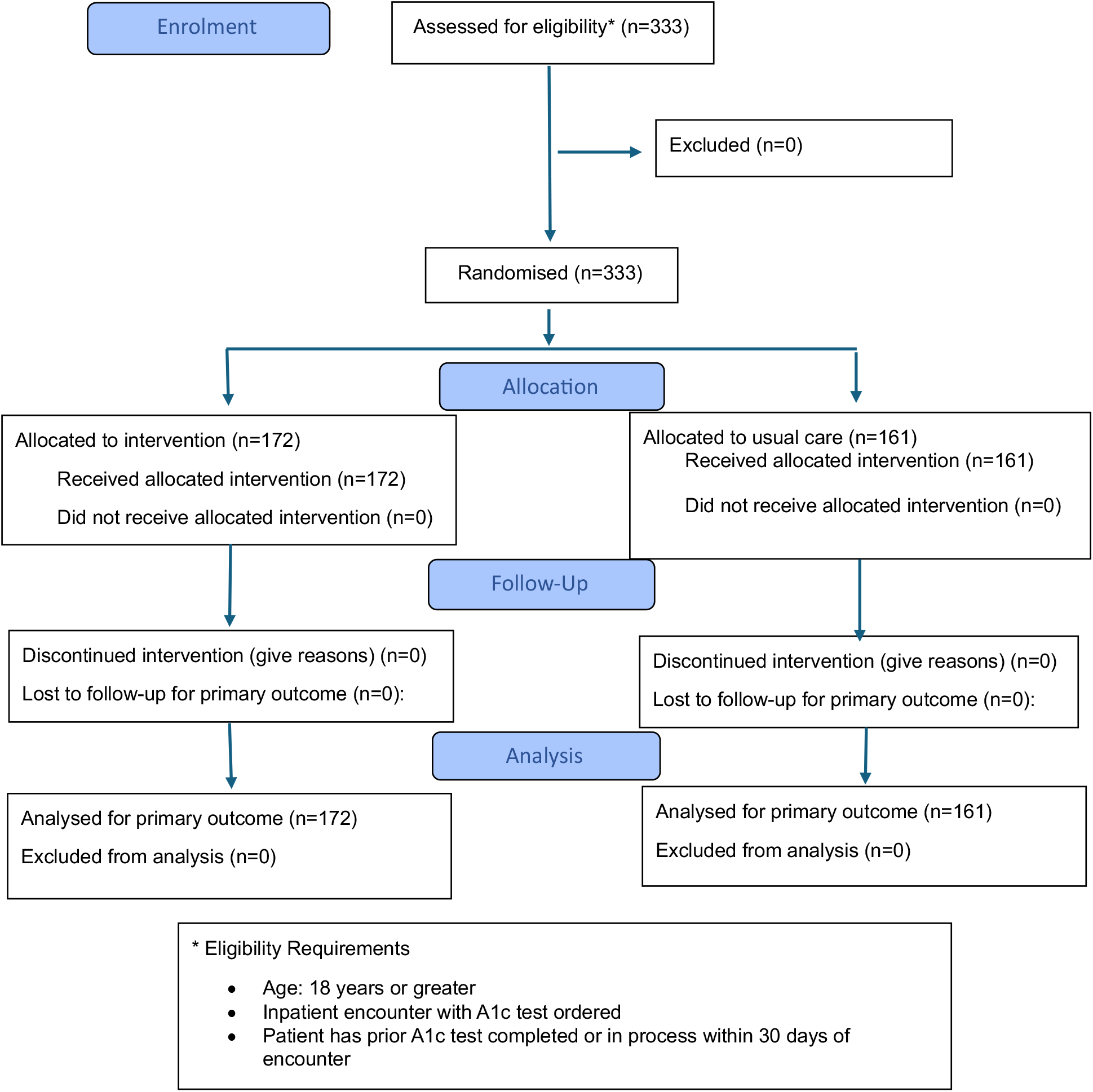
CONSORT 2025 Flow Diagram. Flow diagram of the progress through the phases of a randomised trial of two groups (that is, enrolment, intervention allocation, follow-up, and data analysis)

The total number of orders signed in the intervention arm (primary outcome) was 18 (10%) versus 104 (65%) in the control arm (IRR 0.16, 0.95 CI 0.10 – 0.27). The alert was initially dismissed 34 times, with providers selecting the button “lab still needed,” but in 16 of these 34 encounters the order was never signed, resulting in the total of 18 orders in the intervention arm as reported above. In one case, the provider wrote a free text comment of ““s/p pancreas transplant” as the reason the lab was still needed. Zero orders were cancelled in the control arm. Because the intervention was so effective, when the pre-planned 3-month study period ended, the randomization engine was turned off and 100% of short interval repeat HbA1c tests on adult inpatients now trigger the alert.

In conclusion, our clinical informatics team successfully deployed an innovative point-of-care randomization engine—originally developed at UCSF—into UCLA Health’s EHR, making UCLA Health the second health system in the nation to our knowledge with the capability to instantaneously perform automated block randomization of patients, encounters, clinicians, clinical units, or any other type of “object” at the point-of-care or point-of-order. Our straightforward demostration RCT of a provider-facing alert to reduce low-value repeat lab testing clearly demonstrated the value not only of an integrated randomization system but also of this particular alert as a powerful lever for changing provider behavior. Due to high demand for the POCR randomization engine by providers and researchers at UCLA Health, a new governance council has been established to prioritize resource allocation of this valuable tool.

In summary: The POCR engine is transforming health systems’ ability to conduct pragmatic clinical trials and deliver clinical care within the EHR. marking a critical advancement in the ability to generate high-quality, causal evidence within healthcare systems.

## METHODS

### EHR software vendor

Both UCSF Health and UCLA Health independently manage their own instance of Epic for their main EHR platform. UCSF’s customized implementation of Epic is called APeX (Advancing Patient-Centered Excellence), and UCLA’s customized implementation is called CareConnect.

### Deployment of the POCR engine

The UCLA Health team deployed the UCSF POCR engine following these steps (#1-4 for installation and #5-9 for implementation). See Figure 3:

**Fig. 3.**
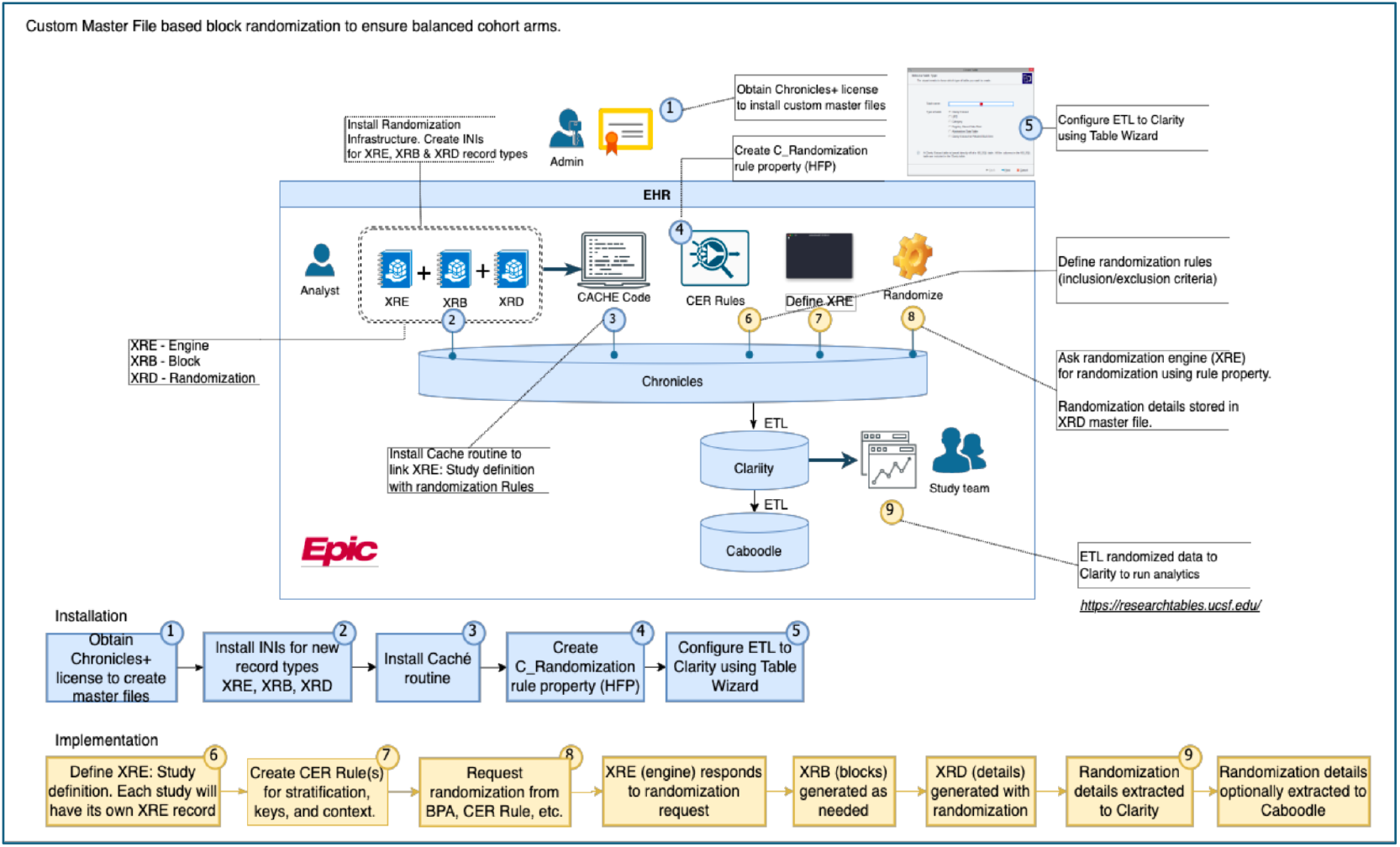
Point of Care Randomization (POCR) Engine. This figure shows the infrastructure required to implement randomization in Epic. Installation steps (blue) are completed once per environment (e.g., testing or production environments) while Implementation steps (yellow) are performed for each randomization project.

1. Purchase Chronicles + license to create master files in Chronicles. (Chronicles is Epic’s proprietary database management system designed specifically to process clinical, operational, and transactional data in real time for every patient encounter.)
2. Install randomization engine infrastructure, creating new Master Files for engine (XRE), block (XRB), and randomization (XRD) types.
3. Install Caché code to utilize the new master files and generate/use randomizations.
4. Configure extraction-transformation-load (ETL) processes to Clarity using Table Wizard.
5. Create randomization rule property (CER rules) for a particular RCT.
6. Define inclusion and exclusion randomization rules and other configuration specifics for a particular RCT.
7. Save configuration details for the RCT in XRE (randomization context, stratification, block size multiplier, etc)
8. Trigger randomization events using rule property; randomization assignments are made according to stratified blocked assignments stored in XRB (new blocks of assignments are generated as needed by the program); and then the randomization assignment and contextual information including strata variables are stored in XRD master file.
9. The randomization assignment is then available for use (immediately and subsequently) as a component of a rule.
10. Data from XRE, XRB, and XRD are automatically extracted to Clarity (a separate relational data warehouse built by Epic for advanced reporting, analytics, and research) and can be optionally extracted to Caboodle (Epic’s enterprise data warehouse).

### Deployment of the randomization engine at UCLA Health

The UCSF AER team provided demonstrations to the UVAS and the UCLA Health ISS and OHIA teams, with step-by-step instructions on implementation of the randomization infrastructure within Epic. The AER team shared specific custom code and were available to answer questions as needed.

### Use-case at UCLA Health: pragmatic RCT of provider-facing alert for low-value repeat HbA1c testing

#### Study Design and Setting

This was a pragmatic, parallel-arm RCT at a large urban academic health system of 3 non-psychiatric adult hospitals with a combined bed count of approximately 1,000.

#### Inclusion Criteria

All inpatients were eligible and silently enrolled if they were age 18 years or older and their provider was attempting to place an order for HbA1c testing within 30 days of a previous HbA1c test result or while results of a recent HbA1c order were still pending.

#### Intervention and Intervention Implementation

The intervention consisted of a provider-facing interruptive alert displaying the date and results of any recent HbA1c testing. Providers had the option either to override the alert and sign their attempted A1c order or to acknowledge the alert and not sign their attempted A1c order.

#### Randomization and Blinding

The UCLA ISS team configured the randomization engine for stratified randomization based on patient age and the value of the most recent HbA1c test. At the instant when a provider attempted to order a HbA1c test for a study-eligible patient, the randomization engine assigned the patient to either the intervention arm or the control arm. Providers and patients were not notified about the study. Providers in the intervention arm were shown the alert but were not aware that it was part of a study or that they had been randomly assigned to an intervention arm.

#### Study Outcomes

The primary outcome was the number of signed HbA1c orders within 30 days of a prior HbA1c result or pending HbA1c order during the observed encounters. Secondary outcomes included frequency of order cancellation and alert overrides in the intervention arm.

#### Data Collection Methods

Data was extracted from UCLA’s Clarity database.

#### Statistical Analyses

The project analyst compared the rate between study arms of our primary outcome – the number of signed HbA1c orders - using Poisson regression. Analyses were performed using Python.

#### Sample Size Analyses

We estimated that the randomization would trigger 32 times per week, with 416 patients enrolled over 3 months and 208 in each study arm, giving us 80% power to detect a 25-percentage point difference in the number of short-interval HbA1c orders with an alpha 0.05.

#### Ethical Issues

The Institutional Review Board (IRB) of the University of California, Los Angeles (UCLA) determined that this quality improvement project was not research involving human subjects as defined by DHHS and FDA regulations and that IRB review and approval were not required. This study was registered with ClinicalTrials.gov (NCT06993805) on April 30, 2025.

## Data Availability

All data produced in the present study are available upon reasonable request to the authors.

## Author Contributions

MP, JP and AG developed the POCR engine and facilitated transfer of technology from UCSF to UCLA, working closely with CS and EC at UCLA. AM, JT and JV led the technical operations at UCLA with supervision from EC, support from WT, and guidance on the clinical aspects of the project from CS, KI and RV. CV conducted data extraction and analyses. SV led on the study design and statistical considerations. CS wrote the first draft of the manuscript, including some text and Figure 3 from a presentation made previously by MP’s UCSF team. Every author read at least one draft of the manuscript, provided substantive edits, and approved the final manuscript.

